# Does anodal tDCS over M1 really enhance motor sequence learning? A non-replication of earlier findings in a double-blind, pre-registered large-sample study in humans

**DOI:** 10.1101/2025.10.06.25337371

**Authors:** Silke Kerstens, Luuk van Boekholdt, Hans Vanderheyden, Louise De Smedt, Nina Seminck, Tine Van Bogaert, Genevieve Albouy, Bradley R. King, Jean-Jacques Orban de Xivry, Myles Mc Laughlin

## Abstract

**Background:** Transcranial direct current stimulation (tDCS) is one of the most widely used noninvasive neuromodulation methods. Despite its popularity, some recent studies highlighted issues about the reproducibility of earlier reported tDCS results. Until recently, it was assumed that tDCS elicits its neuromodulatory effects by increasing cortical excitability through direct polarization of cortical neurons. However, recent studies have shown that the electric field that reaches the cortex is relatively weak, whereas the electric field in the scalp underneath the stimulation electrodes is sufficiently strong to stimulate peripheral nerves, thereby potentially indirectly affecting cortical excitability and plasticity.

**Objective:** In this study, we aimed to replicate the effect of anodal tDCS in enhancing motor sequence learning and investigate if the effect is caused by the polarization of cortical neurons, or more indirectly through stimulation of peripheral nerve in the scalp, or a combination of both mechanisms.

**Methods:** In a double-blind, pre-registered study including 99 healthy young adults, we investigated the effect of 1mA anodal tDCS over the left primary motor cortex (M1) on motor sequence learning in three serial reaction time task (SRTT) sessions using a between-subjects design. In addition to the standard sham condition, we introduced an additional control condition in which the peripheral input was blocked using the BL10 topical anesthetic gel to investigate the potential contribution of peripheral nerve stimulation in mediating tDCS effects in motor sequence learning.

**Results:** Our results provided evidence of motor sequence learning in all three stimulation conditions (p<0.0001). However, no significant differences were observed among the three stimulation conditions (p = 0.94).

**Conclusion:** We were unable to replicate previous findings indicating significant beneficial effects of tDCS on motor sequence learning. Consequently, we were unable to address our main research question of whether tDCS effects are driven by the resulting electric field in the cortex or by stimulation of peripheral nerves in the scalp. This non-replication of one of the presumably most reliable tDCS effects in a much larger sample size than the original studies, among the findings of comparable studies reporting similar outcomes, should prompt a renewed discussion regarding the efficacy of tDCS as a neuromodulation technique, particularly given the earlier reported concerns about its reproducibility and reliability.

## Introduction

Transcranial direct current stimulation (tDCS) is a widely used noninvasive neuromodulation method that involves the application of a low intensity direct current (DC) over scalp electrodes. Early results demonstrated the neuromodulatory potential of the technique (Nitsche and Paulus 2000) sparked a surge of interest among neuroscientists, neurologists, and psychiatrists. Due to its noninvasive nature, low cost, and ease of application compared to other neuromodulation techniques, the field of tDCS has grown rapidly. Despite the large amount of research and numerous studies investigating the clinical applications of the technique, the underlying neurophysiological mechanisms remain poorly understood.

Initially, it was hypothesized that anodal tDCS elicits its neuromodulatory effect by increasing cortical excitability due to depolarization of the resting membrane potential of cortical neurons (Nitsche and Paulus 2000). However, our computational modeling study showed that only a small portion of the applied current effectively reaches the cortex, as most of the current is shunted by highly conductive tissues in between the stimulation electrodes and the targeted cortex such as the scalp, skull, and cerebrospinal fluid (Asamoah, Khatoun, and Mc Laughlin 2019). We demonstrated that when applying 2 mA tDCS, the resulting electric field strength in the cortex is less than 0.5 V/m. Assuming the polarization length constant of excitatory cortical neurons to be 0.2 (Radman et al. 2009), a 1 V/m extracellular electric field should be able to cause a membrane potential shift of 0.2 mV at the soma. This means that the resulting electric field in the cortex of less than 0.5 V/m can only induce a depolarization of the resting membrane potential of up to 0.1 mV. Despite ongoing research on this topic, it remains unclear how such a small shift in the resting membrane potential could account for significant changes in cortical excitability as reported in previous tDCS studies (Nitsche and Paulus 2000).

In an opinion piece, we addressed this question and proposed an alternative mechanism of tDCS, known as the transcutaneous tDCS mechanism (van Boekholdt et al. 2021). In our previously mentioned computational study, we also found that in addition to the unexpectedly weak cortical electric field, the resulting electric field in the scalp underneath the stimulation electrodes can reach levels up to 20 V/m (Asamoah et al. 2019; Rampersad et al. 2014). This is strong enough to elicit action potentials in peripheral nerves that innervate the scalp (So, Stuchly, and Nyenhuis 2004) as evidenced by subjects of tDCS studies often experiencing tingling, burning, or itching sensations during stimulation (Kerstens, Orban de Xivry, and Mc Laughlin 2022; Kessler SK, Turkeltaub PE, Benson JG 2012). In addition to providing input to the somatosensory cortex, stimulation of these cranial and cervical nerves also activates structures in the brainstem where these nerves enter the brain (Fanselow 2012; Mercante et al. 2018) such as the locus coeruleus (LC). As activation of subcortical nodes, such as LC, is known to increase the release of the norepinephrine (NE) and thereby enhance excitability across the cortex, stimulation of peripheral nerves in the scalp during tDCS can also increase cortical excitability, albeit indirectly.

To determine whether the neuromodulatory effects of tDCS are driven by the resulting electric field in the cortex, as originally hypothesized, or by an increase in cortical excitability resulting from stimulation of peripheral nerves in the scalp, it is essential to find a means of isolating and selectively activating the transcranial or the transcutaneous mechanism without engaging the other. We therefore developed a novel tDCS control condition in which the transcutaneous mechanism is blocked by anesthetizing the peripheral nerves in the scalp using the BL10 anesthetic gel (Kerstens et al. 2022).

In this study, we aimed to determine the relative contributions of the transcranial and transcutaneous mechanisms by comparing the effects of regular tDCS, in which both mechanisms are active, with this novel control condition in which only the transcranial mechanism is active. If the observed effects of regular tDCS were to diminish in the control condition, this would indicate that the transcutaneous mechanism plays a significant role in mediating tDCS effects. Conversely, if the effects were to be maintained in the control condition, this would suggest that the transcranial mechanism is primarily responsible for the observed effects of tDCS.

Yet, before being able to evaluate the relative contributions of the neurophysiological mechanisms, our first aim was to replicate the effect of tDCS on motor learning (Buch et al. 2017; Reis et al. 2009). More specifically, we aimed to replicate the effect of anodal tDCS over the primary motor cortex (M1) on motor sequence learning in the serial reaction time task (SRTT) compared to sham stimulation (Dumel et al. 2016; Kang and Paik 2011; Liebrand et al. 2020; Nitsche et al. 2003; Shilo and Lavidor 2019; Stagg et al. 2011). Although the effects of tDCS on motor sequence learning is considered one of the most consistent and reliable effects of tDCS (Buch et al. 2017; Reis et al. 2009) and many studies have demonstrated profound effects (Dumel et al. 2016; Kang and Paik 2011; Liebrand et al. 2020; Nitsche et al. 2003; Shilo and Lavidor 2019; Stagg et al. 2011), it is important to note that not all studies observed significant effect (Horvath, Carter, and Forte 2016) and one study even reported that anodal tDCS over M1 impaired performance on the SRTT (Keitel et al. 2018). Furthermore, among the studies showing significant effects, the timing of tDCS-induced improvements varied considerably, with some occurring during practice and concurrent stimulation (Nitsche et al. 2003; Stagg et al. 2011), some immediately afterward (Dumel et al. 2016), and some only after a 24-hour consolidation following practice (Kang and Paik 2011).

Despite some studies showing these early effects of tDCS either during or immediately after practice, many studies have highlighted that the effect of tDCS on motor learning is more evident after several days of task practice with concurrent stimulation (Dumel et al. 2016; Reis et al. 2009). In fact, a meta-analysis on the effect of multi-session anodal tDCS over M1 on SRTT performance indicated that at least three sessions are required to induce a significant effect of tDCS compared to sham (Hashemirad et al. 2016) when tested in a retest immediately after task practice and stimulation (standardized mean difference: SMD = 0.97), or after a 24h-consolidation interval (SMD = 1.68), with SMD an estimate for the effect size of tDCS compared to sham (Hashemirad et al. 2016).

Accordingly, we aimed to replicate the effect of tDCS on motor sequencing learning in an experimental design including three practice sessions with concurrent stimulation including tests immediately after stimulation, as well as after 24h-consolidation. We hypothesized that anodal tDCS over M1 can significantly improve motor sequence learning performance compared to sham stimulation, when measured in the test immediately after stimulation in the final practice session. Based on this effect, we aimed to test our main hypothesis on relative contribution of the underlying neurophysiological mechanisms by comparing the effect of anodal tDCS to the observed effects of our novel control condition in which the transcutaneous mechanism is blocked.

## Methods

In a large-scale, double-blind, randomized, between-subjects, pre-registered study, we investigated the effect of 1 mA anodal tDCS over left M1 on motor sequence learning. In total, 99 young healthy adults (M: 37%, F: 63%, age: 23 ± 3 years) were included in the study. Upon inclusion, participants were assigned to one of three stimulation groups: anodal tDCS (n_tDCS_ = 33) group, the novel control condition in which the transcutaneous tDCS mechanism was blocked using the BL10 anesthetic gel(Kerstens et al. 2022), (n_tDCS+BL10_ = 33), or the sham control (n_sham_ = 33) group.

### Ethical approval

This study was approved by the UZ/KU Leuven Research Ethical Committee (S63709) and performed in accordance with the ethical principles outlined in the Declaration of Helsinki. The study was registered on ClinicalTrials.gov (NCT04577677) and preregistered on the <u>Open Science Framework</u>. Prior to preregistration, a pilot study including twelve subjects was performed to optimize the motor sequence learning paradigm and the tDCS stimulation parameters.

### Subjects

An a-priori power analysis (95% power, α = 0.05, two-sided) indicated a minimum of 33 subjects per group to detect tDCS effects on motor sequence learning compared to sham, yielding a total sample size of 99 subjects. All subjects met standard inclusion and exclusion criteria for tDCS motor learning studies and were aged 18–40, right-handed, in good general health, and free from neurological, psychiatric, or musculoskeletal conditions, as well as contraindications to brain stimulation, and lidocaine or benzocaine allergies. Individuals with prior SRTT experience, musical training, gaming or typing expertise, metal implants or piercings near stimulation sites, medication use, or potential pregnancy were excluded. All participants provided written informed consent and followed pre-session guidelines, including adequate rest and abstaining from stimulating substances.

### Stimulation conditions

In the anodal tDCS condition, 1 mA anodal tDCS was applied over left M1 (C3) using the Signa-gel filled 3D-printable cup electrodes (Kerstens et al. 2025) with a diameter of 19 mm and the neuroConn DC-Stimulator Plus (neurocare Group, Munich, Germany) using Ag/AgCl connector cables. The return electrode was located in the contralateral supraorbital position (FP2), as illustrated in Figure 1B. Prior to electrode placement, the scalp was cleaned to ensure optimal conductivity. Finally, a conductivity test was performed to confirm that the system’s impedance was below 5 kΩ before stimulation was started with a 1-second ramp to 1 mA.

**Figure 1.**
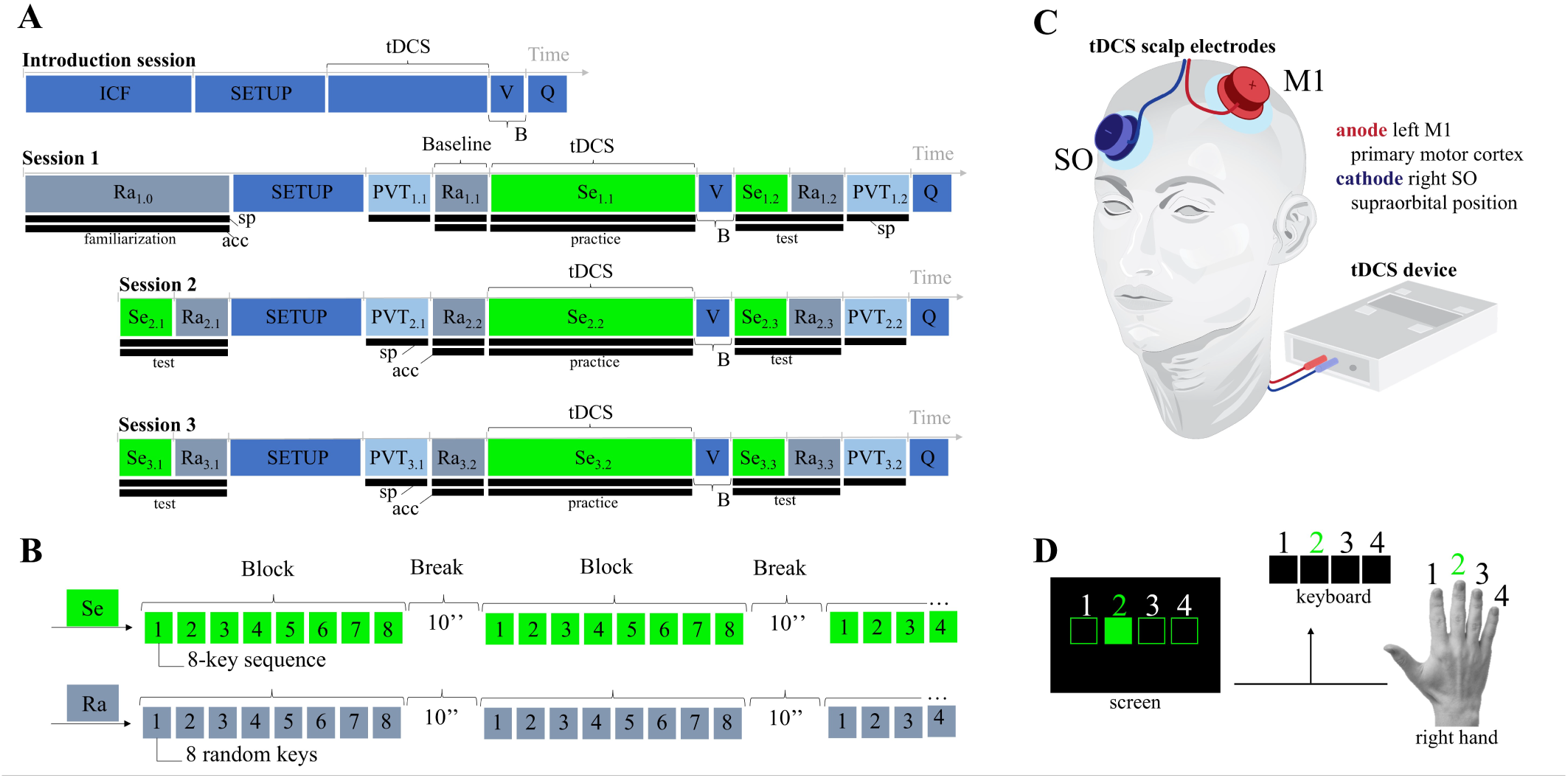
Study protocol. **A)** The study protocol consisted of an introduction session and three motor sequence learning sessions. In the introduction session, subjects were informed about the study in detail and informed consent was obtained. Next, stimulation was applied for 10 minutes to get acquainted with the stimulation. At the start of session 1, subjects familiarized with the task in 16 random (Ra) blocks. Next, the tDCS stimulation equipment was applied and subjects before subjects performed the PVT (no stimulation). Subjects then performed four random (Ra) blocks to establish a baseline performance for the SRTT. Subsequently, subjects performed a full run of 16 sequenced (Se) SRTT blocks, during which stimulation was applied continuously with the duration of stimulation depending on the time needed to complete the task. After completing the sequential SRTT, subjects were given a brief two-minute break (B), during which they were asked to indicate their stimulation sensation on a visual analogue scale (V). Next, motor performance on both the sequential and the random SRTT was tested with four sequenced (Se) blocks and four random (Ra) blocks, respectively. Immediately after this test, the subjects performed a second PVT to allow a comparison of the subjects’ performance on the PVT before and after stimulation and assess the effect of task practice and stimulation on vigilance. Finally, at the end of each session, subjects reported on potential stimulation side effects in a short questionnaire (Q). Session 2 and session 3 were similar to session 1, with only one major difference at the start of the sessions. Instead of the familiarization in the beginning of session 1, an additional test was included in the beginning of session 2 and session 3 to assess the effect of 24h-consolidation interval on motor memories. During the SRTT, both speed (sp) and accuracy (acc) were measured to assess performance. During the PVT only reaction times in terms of speed (sp) were recorded. **B)** Each practice block in the SRTT consisted of 64 key presses. For the sequenced (Se) blocks, this involved 8 repetitions of the 8-element sequence. For the random blocks (Ra), this involved 64 pseudo-randomized key presses. Between each block, subjects had a brief rest period of 10 seconds. **C)** Stimulation was applied over Signa gel-filled cup electrodes with the anode located over the left primary motor cortex (M1) and the return electrode in the contralateral supraorbital (SO) position with the other stimulation parameters depending on the stimulation condition. **D)** During the SRTTs, four empty squares were represented horizontally on a screen and corresponded to four keys on a keyboard, and to the four fingers of the right hand (excluding the thumb). During the task, the visual stimuli were presented by filling one of the squares while subjects were instructed to respond to the stimuli as fast and as accurately as possible by pressing the corresponding key with the corresponding finger. Upon a key press, either correct or incorrect, the next stimulus was presented on the screen immediately. Each block consisted of 64 key presses and rest periods between practice blocks were visually signaled by the squares turning red. To allow refocusing for the next block, the squares transitioned back to green two seconds before the next stimulus was represented on the screen.

For the sham condition, the same electrode setup was used, and a brief stimulation was applied to mimic the sensations of tDCS before it was ramped down again after ten seconds, thereby controlling for potential bias induced by subject’s expectation related to the effects of tDCS. In the novel control condition, again the same stimulation electrode setup was used. However, to block the transcutaneous tDCS mechanism, an additional layer of the anesthetic BL10 gel (Kerstens et al. 2022), containing 5% lidocaine and 5% benzocaine, was applied on the scalp underneath the cup electrodes precisely matching the electrode’s location and surface area to keep the stimulation surface area consistent.

### Motor sequence learning

The motor sequence learning paradigm comprised an introduction session followed by three motor sequence learning sessions conducted on three consecutive days (Figure 1A). During the introduction session, participants received detailed study information, provided written informed consent, and underwent 10 minutes of tDCS to familiarize themselves with the stimulation prior to the practice sessions. In each motor sequence learning session, participants performed a sequenced serial reaction time task (Van Roy et al. 2024) (SRTT) while receiving concurrent stimulation (Figure 1C). Each SRTT consisted of sixteen sequenced blocks (Se) as indicated in Figure 1A: Se_1.1_, Se_2.2_, and Se_3.2_, with each block comprising 8 repetitions of an 8-element stimuli (Figure 1B), with 10-second rest intervals between each block to reduce fatigue (Figure 1B).

At the start of each session, subjects were instructed on how to perform the SRTT. Four empty squares were displayed on a screen, each corresponding to a key on the keyboard, and a finger of the right hand (Figure 1D). Visual stimuli were presented by filling one the squares (Figure 1D) and subjects were asked to press the correct key with the corresponding finger as quickly and accurately as possible. After each key press, whether correct or incorrect, the next stimulus was immediately presented.

#### Serial reaction time task sequences

Each subject was assigned to practice one of the following 8-element sequences: sequence A: 2-4-1-3-2-4-3-1, sequence B: 1-3-1-4-3-2-4-2, or sequence C: 3-1-4-2-1-3-2-4, with 1 referring to (from left to right) the first square on the screen and the first key on the keyboard, and the right index finger, et cetera. To avoid bias from specific motor sequences, subjects were randomly assigned to one of the sequences, ensuring equal distribution across the three stimulation conditions.

#### Serial reaction time task familiarization

At the beginning of the first session, an additional familiarization phase was implemented, consisting of 16 random blocks (Ra) of 64 stimuli each (Figure 1A: Ra_1.0_), in which the squares were filled in a pseudorandomized order. These blocks allowed familiarization and controlled for non–sequence-specific visuomotor habituation, with no stimulation applied. In addition, four random blocks were included at the beginning of each session (Figure 1A: Ra_1.1_, Ra_2.2_, Ra_3.2_ to familiarize subjects before the sequenced SRTT and thereby minimize initial performance effects.

#### Baseline performance

The four additional random blocks performed after task familiarization (Figure 1A: Ra_1.0_) and before the start of the first sequenced SRTT in session 1, were defined as baseline performance (Figure 1A: Ra_1.1_).

#### Motor sequence learning improvements across sessions

Immediately after each sequenced SRTT (Figure 1A: Se_1.1_, Se_2.2_, and Se_3.2)_, potential improvements in motor sequence learning performance were assessed in a test by comparing the subject’s performance in four sequenced blocks (Figure 1A: Se_1.2_, Se_2.3_, and Se_3.3_) to their performance in four random blocks (Figure 1A: Ra_1.2_, Ra_2.3_, and Ra_3.3_), during which no stimulation was applied.

#### Consolidation effect on motor sequence memory

Finally, to assess the effect of 24h consolidation on motor memory, additional tests were added at the start of sessions 2 and 3 (Figure 1A: Se_2.1_ and Ra_2.1_, Se_3.1_ and Ra_3.1_).

### Psychomotor vigilance task

In addition to the motor sequence learning paradigm, subjects also performed two psychomotor vigilance tasks (PVT) each session: one at the start (Figure 1A: PVT_i.1_ with i = 1,2, and 3) and one at the end (Figure 1A: PVT_i.1_ with i = 1,2, and 3) of each session. PVT is a widely used reaction time task to assess an individual’s sustained attention, alertness and vigilance allowing evaluation of whether motor sequence learning sessions or concurrent stimulation affected subject’s alertness or induced significant changes in fatigue. During the PVT, 30 visual stimuli were presented on a screen at random intervals of 2–10 seconds. Subjects were instructed to respond as quickly as possible to each stimulus by pressing a key with their right index finger immediately after detecting the visual cue. This shorter version(Matsangas and Shattuck 2018) of the original 100 trial PVT (Dinges and Powell 1985) has also been shown to reliably assess sustained vigilance (Basner and Rubinstein 2011; Loh et al. 2004).

### Stimulation sensation

The assess the effectiveness of the anesthesia in blocking the peripheral nerves in the scalp (Kerstens et al. 2022), subjects were asked to indicate the intensity of the stimulation sensation at the end of each session based on a visual analog scale (VAS) from 0 to 100 with intermediate levels: 0 no sensation, 25 uncomfortable, 50 very uncomfortable, 75 painful, 100 very painful.

### Side effects

Subjects were also asked to report if they experienced any of the following tDCS-related side effects (Brunoni et al. 2011; Buch et al. 2017; Kerstens et al. 2022; Kessler SK, Turkeltaub PE, Benson JG 2012; Poreisz et al. 2007): tingling, pinching, itchiness, warm or even slightly burning sensations in the scalp, pressure underneath the stimulation electrodes, headaches, dizziness, nausea, phosphenes, metallic taste in the mouth, general discomfort, mild pain, or fatigue, and to rate their intensity using the following levels: very slightly, slightly, moderately, considerably, strongly, or very strongly.

Finally, they indicated the timing of each reported side effect as: only at the start, only at the end, once during stimulation, at multiple occasions, or consistently throughout stimulation.

## Data analysis

### Motor sequence learning

The motor sequence learning paradigm was designed to facilitate motor sequence learning over time. Improvements were assessed based on changes in the mean reaction times (RTs), defined as the time between stimulus presentation and key press response in milliseconds (ms), per block. To avoid bias from incorrect responses, only RTs from correct responses were included when averaging per block. As described in the preregistration, subjects who showed no improvement across practice sessions were considered noncompliant or disengaged and were excluded from further analysis. Additionally, overall accuracy was calculated for each subject as the mean across sessions to assess task engagement, and subjects with accuracy below 85% were also excluded. All analyses were conducted in MATLAB 2019 (MathWorks® R2019b).

#### Baseline performance

Baseline performance for each subject was calculated as the mean reaction time across the four random blocks in Ra_1.1_, with group-specific baselines obtained by averaging these values across all subjects within each stimulation condition. Baseline performance was compared between stimulation conditions using unpaired two-sample t-tests, with the significance threshold adjusted for multiple comparisons using a Bonferroni correction to α = 0.05/3.

#### Motor sequence learning improvements across sessions

Improvements in SRTT performance across sessions were assessed based on the reduction in mean RTs in the four sequenced blocks of the tests at the end of each session (Se_1.2,_ Se_2.3,_ and Se_3.3_) relative to baseline. For each stimulation condition respectively, improvements across sessions were evaluated by comparing their performance each session to baseline using unpaired two-sample t-tests, with the significance level adjusted for multiple comparisons using a Bonferroni correction to α = 0.05/3.

#### Effect of stimulation conditions on motor sequence learning measured immediately after stimulation

At the end of each practice session, improvements in motor sequence learning were assessed immediately after stimulation as the difference between the mean reaction times of the four random blocks and the four sequenced blocks in the test at the end of each session. Specifically, motor sequence learning after practice session 1 was calculated as mean(Ra_1.2_) – mean(Se_1.2_), after session 2 as mean(Ra_2.3_) – mean(Se_2.3_), and after session 3 as mean(Ra_3.3_) – mean(Se_3.3_). The effect of stimulation condition on motor sequence learning immediately after stimulation across sessions was then analyzed using a linear mixed model (LMM): motor sequence learning immediately after stimulation ∼ 1 + session + stimulation condition + session × stimulation condition + (1 | subject), with fixed effects for session and stimulation condition and a random intercept for subject. To further examine differences between stimulation conditions within each session, unpaired two-sample t-tests were conducted. To correct for multiple comparisons across the three stimulation conditions, the significance level was adjusted using a Bonferroni correction to α = 0.05/3. Finally, a Bayes factor was calculated to assess the likelihood of the null hypothesis that tDCS does not enhance motor sequence learning after three practice sessions compared to sham with respect to the alternative hypothesis that tDCS does enhance motor sequence learning.

#### Effect of stimulation conditions on online motor sequence learning

In addition to improvements across sessions, the effect of stimulation conditions on online motor sequence learning during concurrent stimulation was analyzed for each session respectively by comparing the mean reaction times per block over time across stimulation conditions. For each session, block-wise mean RTs were averaged across subjects within each stimulation condition to obtain group-level mean RTs. To ensure comparability between conditions, the group-level mean RTs were baseline-corrected using the group-specific baselines. For each session, the effect of stimulation on online motor sequence learning was analyzed using a repeated-measures ANOVA, with blocks as the within-subject factor. Potential differences between stimulation conditions were then examined for each block by pairwise comparisons of the mean reaction time per condition using unpaired two-sample t-tests, with a Bonferroni-corrected significance threshold of α = 0.05/3.

#### Consolidation effect on motor sequence memory

The effect of 24-hours overnight consolidation on motor sequence memory was assessed by comparing performance in the tests at the end of sessions 1 and 2 with the retests 24h later at the beginning of sessions 2 and 3 respectively, using unpaired two-sample t-tests, with a Bonferroni-corrected significance threshold of α = 0.05/3.

#### Serial reaction time task sequences

To assess potential differences in motor sequence learning difficulty across SRTT sequences, the LMM on effects measured immediately after stimulation was refitted with ‘SRTT sequence’ added as a fixed effect: ‘motor sequence learning motor immediately after stimulation ∼ 1 + session + stimulation condition + SRTT sequence + session x stimulation condition + session x SRTT sequence + stimulation condition x SRTT sequence + (1 | subject) and verified whether the sequences significantly influenced the observed results.

### Psychomotor vigilance task

During the PVT, response times, defined as the time intervals between stimulus presentation and the key press responses, were recorded for each trial. To assess whether motor sequence learning or concurrent stimulation affected vigilance, PVT performance was compared before and after practice with concurrent stimulation using the following distribution features: median reaction time, interquartile range, skewness, and kurtosis. Higher median RTs, larger interquartile ranges, and higher skewness or kurtosis indicated lower vigilance and vice versa. For each session and stimulation condition, potential changes in these features were analyzed using unpaired two-sample t-tests.

### Stimulation sensations

To analyze the self-reported stimulation sensations, the mean rating per stimulation condition was calculated for each session and compared using unpaired two-sample t-tests, with the significance threshold Bonferroni-corrected to 0.05/3 for multiple comparisons.

### Side effects

To enable statistical analyses of reported side effect intensities, the intensity levels were quantified on a scale (1: very slightly, 2: slightly, 3: moderately, 4: considerably, 5: strongly, 6: very strongly) and averaged per side effect, session, and stimulation condition. For the most frequently reported side effects, the effects of ‘stimulation condition’ (between-subject factor) and ‘session’ (within-subject factor) on intensity were assessed using ANOVA, followed by more in depth post-hoc testing for side effects showing a significant main effect of stimulation condition. Finally, the occurrence of side effects was analyzed by counting the number of subjects reporting them at specific time points.

## Results

Our results demonstrated that all stimulation groups showed sustained improvements in motor sequence learning across sessions, as well as significant online learning during practice with concurrent stimulation in each session. However, no significant differences were observed between the stimulation groups, not immediately following a single session, or after multiple sessions, during practice and concurrent stimulation, or in measures of consolidation.

### Motor sequence learning

#### Baseline performance

After the initial familiarization phase in the first session, all subjects demonstrated comparable baseline performance (Figure 2) across stimulation conditions (mean RT: meanRT_tDCs_ = 406 ± 33 ms, meanRT_tDCs+BL10_ = 397 ± 38 ms, and meanRT_sham_ = 398 ± 35 ms) with no significant differences between them (p_tDCS – Sham_ = 0.474, p_tDCS – tDCS+BL10_ = 0.342, and p_tDCS+BL10 – sham_ = 0.785).

**Figure 2.**
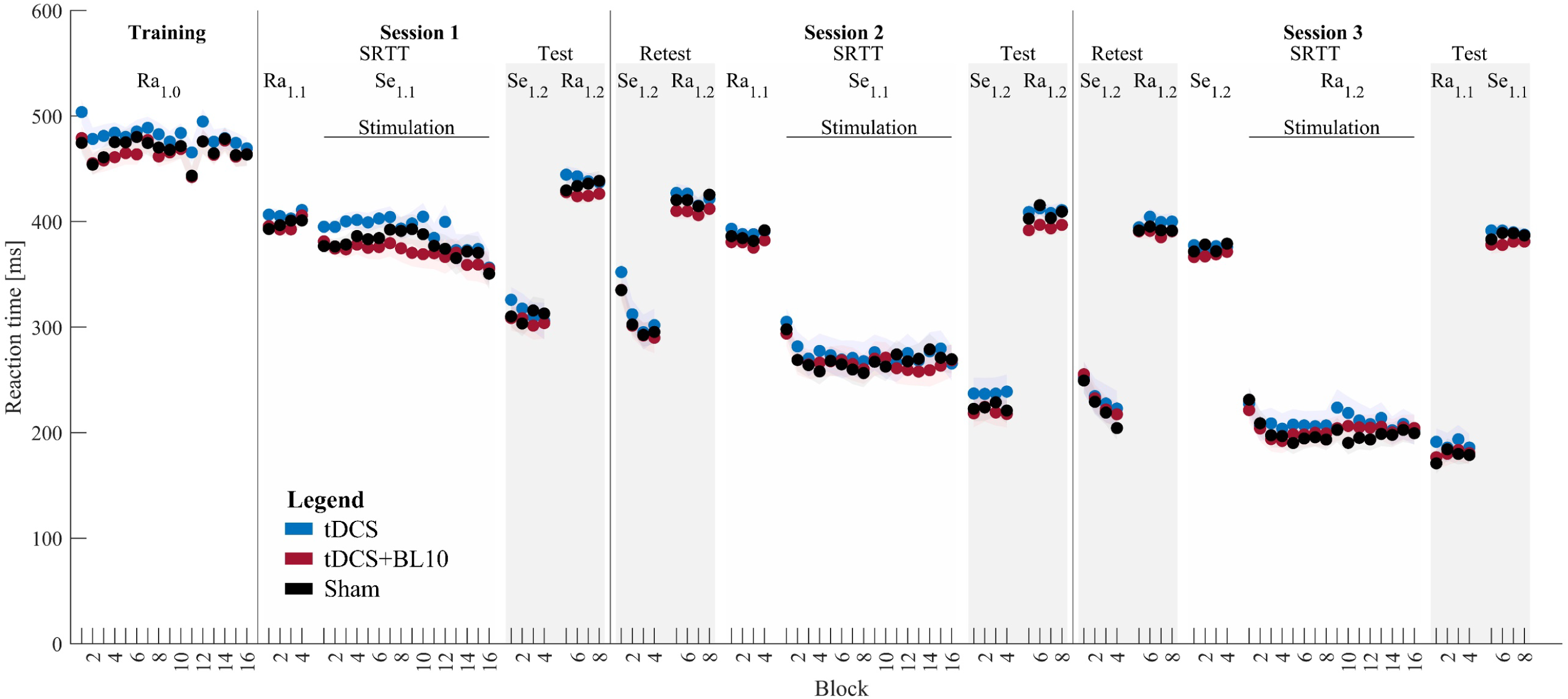
**Motor sequence learning paradigm results**. Mean reaction times (RTs) per block are shown in chronological order, beginning with the familiarization phase, followed by the first sequenced SRTT and subsequent test at the end of session 1. Sessions 2 and 3 each began with a retest, continued with a sequenced SRTT, and ended each with a final test. Data are displayed for tDCS (blue), tDCS+BL10 (red), and sham stimulation (black). Shaded gradients represent 95% confidence intervals, calculated as the standard deviation divided by the square root of the number of subjects per stimulation group.

#### Motor sequence learning improvements across sessions

Significant improvements in motor sequence learning were observed for all stimulation conditions across all sessions, as evidenced by the significant reductions in mean RT in the sequenced blocks of the tests at the end of each session (p < 0.001), with the exception of session 3, where the tDCS (p = 0.026) and tDCS+BL10 (p = 0.023) conditions did not reach significance after correction for multiple comparisons. As the design of the paradigm inherently facilitated motor sequence learning, it is not surprising that their performance improved over time (Figure 2). In fact, individuals who did not show any improvement after three practice sessions were considered to lack task compliance or engagement and were therefore excluded from further data analysis. This was the case for 8 subjects in total, of which 5 subjects in the tDCS group, and 3 subjects in the sham group. No further subjects were excluded based on low accuracy, as all remaining subjects achieved an overall accuracy higher than 85%.

Improvements over time should therefore not be interpreted as a standalone result but primarily reflect adherence to task requirements. Of greater interest are the progressive improvements in motor learning observed across sessions, reflecting ongoing learning with practice (Figure 3). Figure 3B illustrates individual subject performance for each session relative to baseline and to previous sessions, with each dot below the diagonal indicating an improvement in performance. Only a few dots above the diagonal in the plots comparing session 3 to session 2 indicate that a small number of subjects did not show improvement in the final session. Although the observed improvements at the group level in session 3 relative to session 2 did not reach significance for the tDCS (p = 0.026) and tDCS+BL10 (p = 0.023) conditions after correction of the significance level for multiple comparisons, they represent an important result for further discussion in this paper: rather than plateauing early in the paradigm, subjects exhibited continued motor sequence learning across successive sessions.

**Figure 3.**
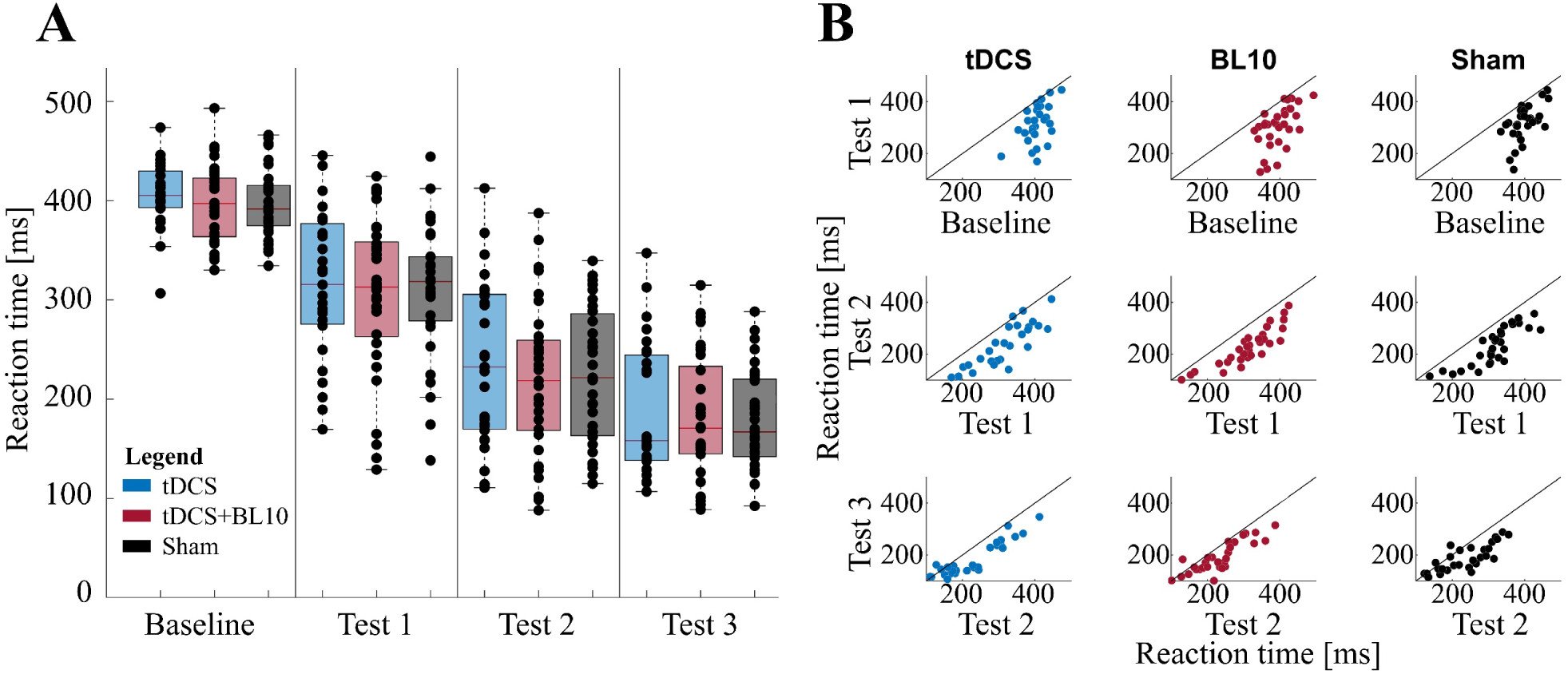
**Motor sequence learning improvements across sessions**. (**A**) Box plots show significant reductions in reaction time at the end of each session compared to baseline and the preceding session across all stimulation conditions. In each box plot, the red line marks the median, the box edges indicate the 25th and 75th percentiles, and whiskers extend to the most extreme values not considered outliers. Dots represent individual participants: tDCS (n = 28, blue), tDCS+BL10 (n = 33, red), and sham (n = 30, black). (**B**) Scatter plots show mean reaction times for individual participants in the sequenced blocks in test 1 versus baseline (top row), test 2 versus test 1 (middle row), and test 3 versus test 2 (bottom row) to evaluate session-to-session improvements. Dots below the diagonal (black line) indicate faster reaction times (improved performance) relative to the previous test or baseline, consistent with motor sequence learning, while dots above the line indicate slower reaction times.

#### Effect of stimulation conditions on motor sequence learning measured immediately after stimulation

Participants in all stimulation conditions showed significant learning improvements across sessions (Figure 4) as indicated by the LMM main effect of session: p < 0.001. However, no significant differences were found between stimulation conditions (LMM main effect of stimulation condition: p = 0.939). Unpaired two-sample t-tests comparing improvements in motor sequence learning between stimulation conditions immediately after the first practice session revealed no significant differences: between the tDCS and sham groups (p_tDCS – sham_ = 0.711, Cohen’s d_tDCS – sham_ = 0.10), between tDCS+BL10 and sham (p_tDCS+BL10 – sham_ = 0.929, Cohen’s d_tDCS+BL10 – sham_ = 0.02), or between tDCS and tDCS+(p_tDCS – tDCS+BL10_ = 0.802, Cohen’s d_tDCS+BL10 – sham_ = 0.06). At the end of session 2, no significant differences were again observed: tDCS versus sham (p_tDCS – sham_ = 0.827, Cohen’s _dtDCS – sham_ = 0.06), tDCS+BL10 versus sham (p _tDCS+BL10 – sham_ = 0.795, Cohen’s d_tDCS+BL10 – sham_ = 0.07) and tDCS versus tDCS+BL10 (p_tDCS – tDCS+BL10_ = 0.987, Cohen’s d_tDCS – tDCS+BL10_ < 0.01). Session 3 yielded similar results, with no significant differences between tDCS and sham (p_tDCS – Sham_ = 0.717, Cohen’s d_tDCS – sham_ = 0.10), tDCS+BL10 and sham (p _tDCS+BL10 – sham_ = 0.529, Cohen’s d _tDCS+BL10 – sham_ = 0.16), or tDCS and tDCS+BL10 (p_tDCS – tDCS+BL10_ = 0.847, Cohen’s d_tDCS – tDCS+BL10_ = 0.05). These findings were further supported by the Bayes factor (Bayes_tDCS – sham_ = 0.153) being well below 1, providing substantial evidence against the hypothesis that tDCS can enhance motor sequence learning, and supporting the conclusion that tDCS had little to no effect compared to sham after three practice sessions.

**Figure 4.**
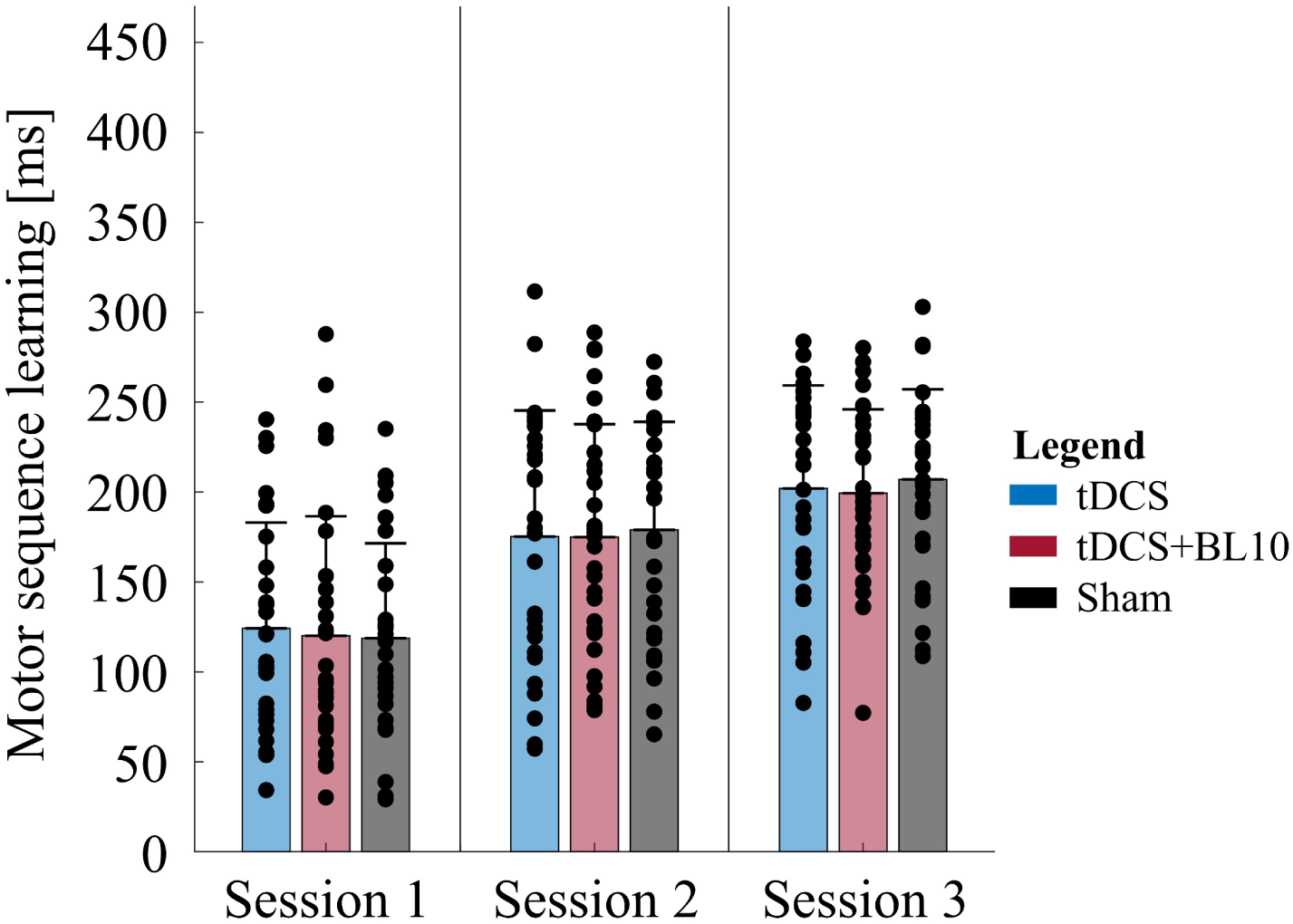
**Effect of stimulation conditions on motor sequence learning measured immediately after stimulation**. Motor sequence learning improvements were calculated as the difference between the mean reaction times of four sequenced blocks and four random blocks in the test at the end of each session. The bar plots represent group means and show comparable learning levels across the three stimulation conditions: tDCS (n = 28, blue), tDCS+BL10 (n = 33, red), and sham (n = 30, black), with the error bars indicate standard deviations.

#### Effect of stimulation conditions on online motor sequence learning

In each session, all stimulation groups showed significant improvements in online motor sequence learning (effect of block in repeated-measures ANOVA: p < 0.001 in all sessions) during concurrent stimulation (Figure 5). However, no effect of stimulation conditions was observed in any session (effect of stimulation condition in repeated-measures ANOVA: session 1: p = 0.488, session 2: p = 0.564, session 3: p = 0.409), and pairwise comparisons between stimulation conditions for each block respectively revealed no significant differences either.

**Figure 5.**
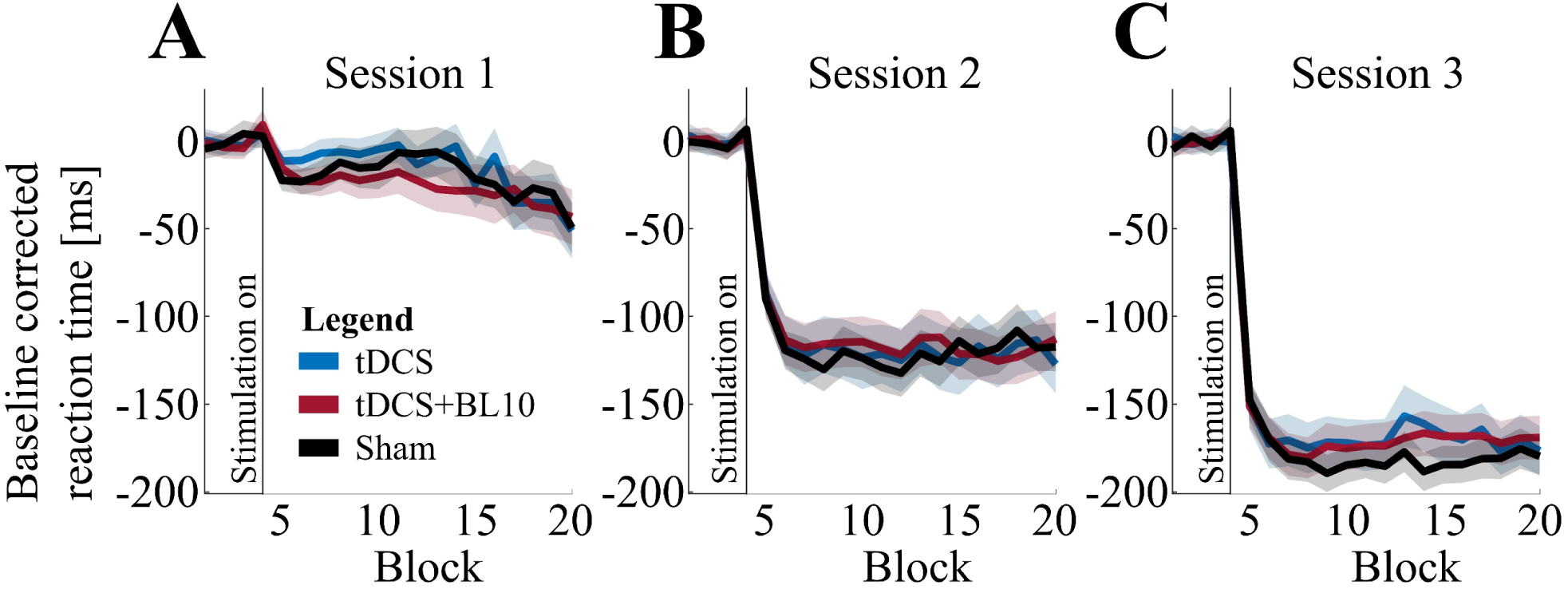
Effect of stimulation conditions on online motor sequence learning. Baseline-corrected online motor sequence learning curves in (A) session 1, (B) session 2, and (C) session 3 for each stimulation condition: tDCS (n_tDCS_ = 28, blue), tDCS+BL10 (n_tDCS+BL10_ = 33, red), and sham (n_sham_ = 30, black), including the baseline-corrected four random blocks preceding each sequenced SRTT. Stimulation was applied only during practice on the sequenced SRTTs, with onset indicated by the vertical black lines. Shaded areas represent 95% confidence intervals (0.95 × SD / √n, with n number of subjects per stimulation condition) around the mean RTs for each stimulation condition. Across all sessions, learning curves were very similar between stimulation conditions.

#### Consolidation effect on motor sequence memory

Comparison of motor sequence learning at the end of each session with performance at the beginning of the subsequent session revealed that improvements were maintained but did not further increase following 24-hour overnight consolidation (Figure 6), with no significant differences between the stimulation conditions.

**Figure 6.**
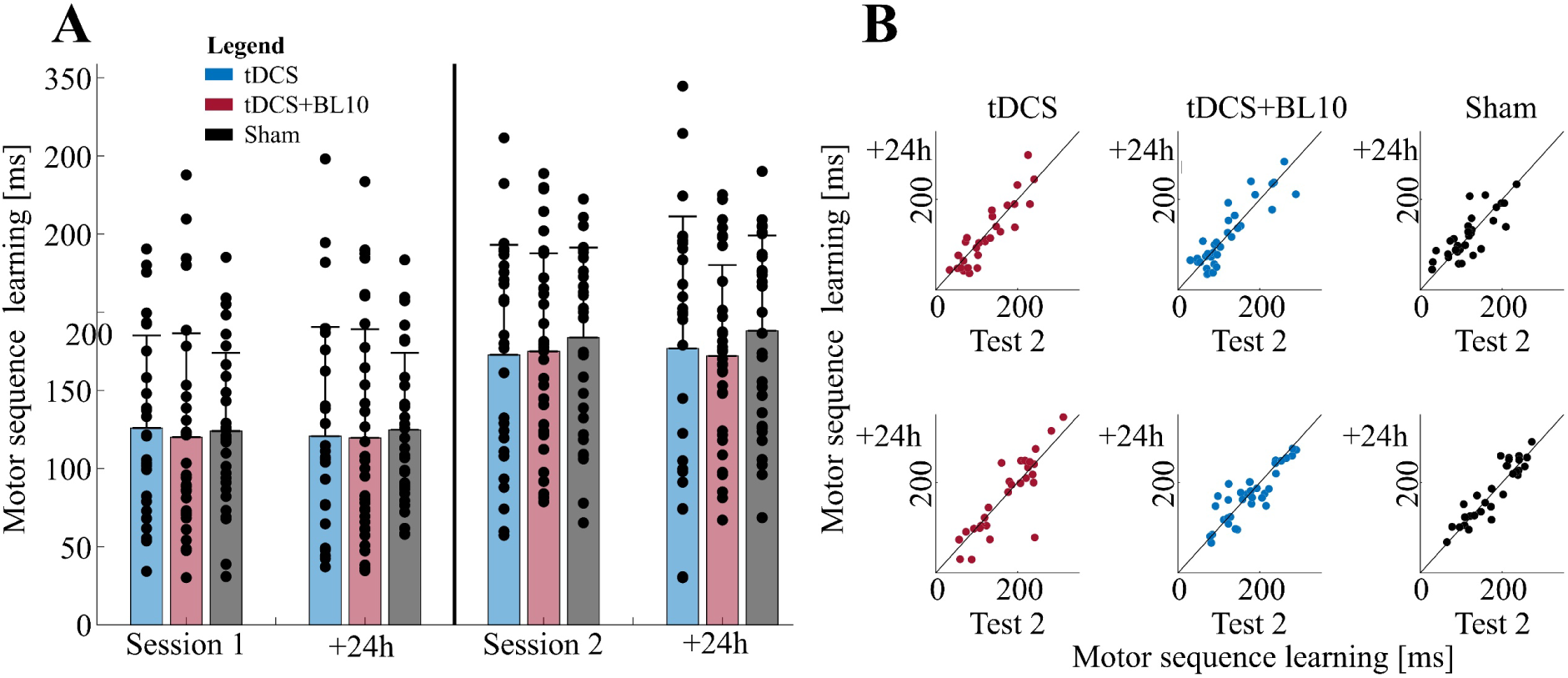
Consolidation effect on motor sequence memory. **A**) Bar plots show motor sequence learning immediately after stimulation in session 1 and 2, as well as after 24-hour consolidation (+24h) for tDCS (n_tDCS_ = 28, blue), tDCS+BL10 (n_tDCS+BL10_ = 33, red) and sham (n_sham_ = 30, black). Bars represent group means, with error bars indicating standard deviations. **B**) Scatter plots show individual subject performance, comparing motor sequence learning in the test at the end of session 1 (first row) and 2 (second row), compared to after 24-hours consolidation. Dots below the diagonal (black line) indicate improvement after consolidation, while dots above indicate no improvement with t-test statistics showing no enhancements after 24h consolidation.

#### Serial reaction time task sequences

Even after excluding non-compliant subjects, the SRTT sequences remained evenly distributed both across stimulation conditions (sequence A: tDCS 31%, tDCS+BL10 33%, sham 36%, sequence B: tDCS 30%, tDCS+BL10 37%, sham 33%, and sequence C: tDCS 29%, tDCS+BL10 39%, sham 32%) and within each condition (tDCS: A 37%, B 33%, C 30%, tDCS+BL10: A 33%, B 33%, C 33%, and sham: A 39%, B 32%, C 29%). Moreover, the LMM including ‘SRTT sequence’ as an additional fixed effect revealed no significant main effect of sequence (LMM main effect of SRTT sequence: p = 0.888), indicating that all employed sequences posed an equivalent level of learning difficulty.

### Psychomotor vigilance task

The PVT results indicated no effect of task practice or stimulation on vigilance, with no significant changes observed when comparing PVT measures before versus after stimulation (Figure 7). For all stimulation conditions, median reaction times did not differ significantly before versus after stimulation (unpaired two-sample t-test: p_tDCS_ > 0.05, p _tDCS+BL10_ > 0.05, p_sham_ > 0.05). Similarly, the interquartile range showed no significant differences for any condition (p_tDCS_ > 0.05, p _tDCS+BL10_ > 0.05, p_sham_ > 0.05). Finally, skewness and kurtosis of the reaction time distributions were slightly higher after tDCS, but these differences were not statistically significant for any stimulation condition (Skewness: p_tDCS_ > 0.05, p _tDCS+BL10_ > 0.05, p_sham_ > 0.05, and Kurtosis: p_tDCS_ > 0.05, p _tDCS+BL10_ > 0.05, p_sham_ > 0.05).

**Figure 7.**
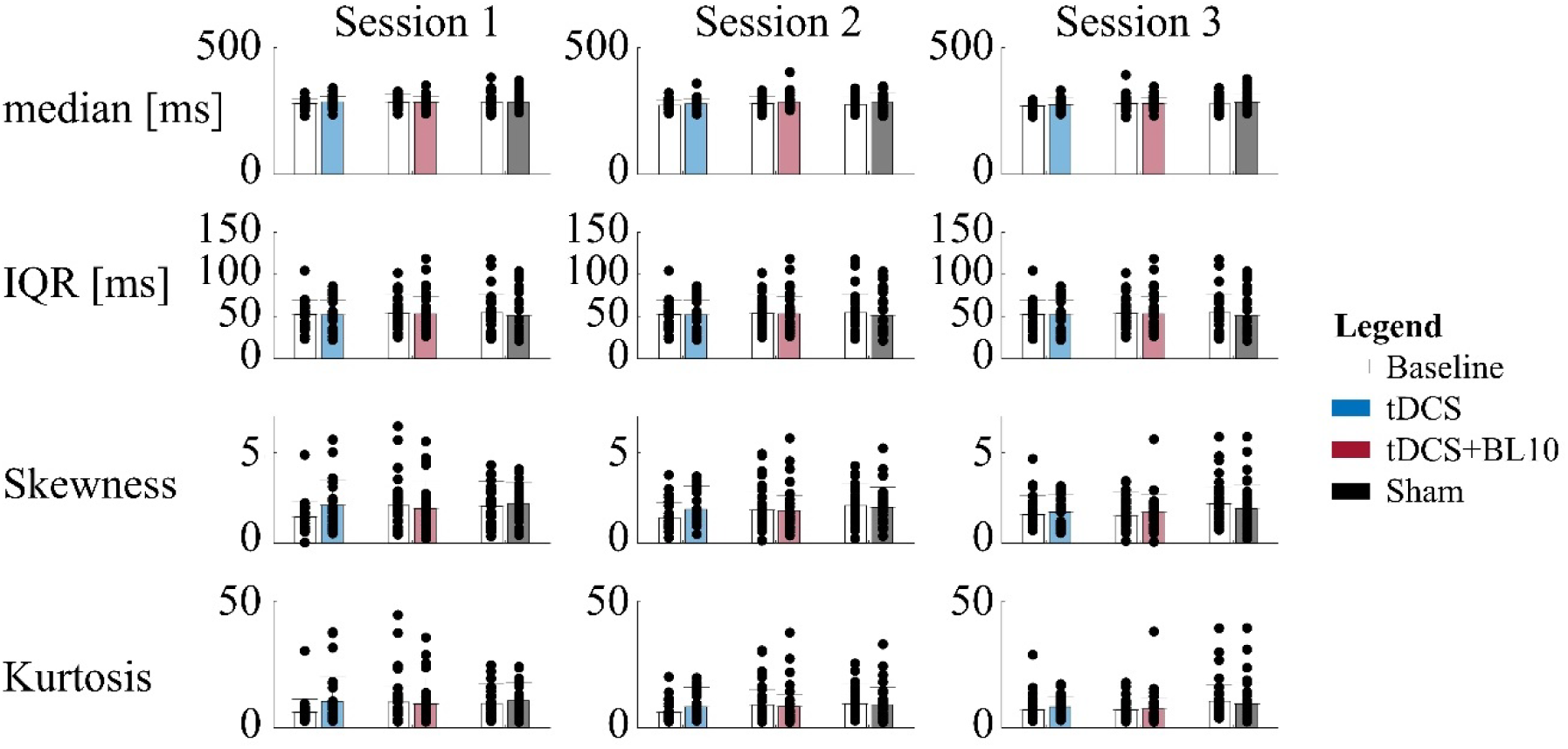
**Psychomotor vigilance task results**. The subplots in the figure show PVT measures of vigilance per session (columns): median reaction time (first row), interquartile range (IQR, second row), skewness (third row), and kurtosis (fourth row). Data are shown per stimulation condition: tDCS (n_tDCS_ = 28, blue), tDCS+BL10 (n_tDCS+BL10_ = 33, red), and sham (n_sham_ = 30, black), with the white bars representing baseline values before practice and concurrent stimulation. No significant changes were observed between pre-and post-stimulation measures, indicating that stimulation had no effect on vigilance in any of the stimulation conditions.

### Stimulation sensations

Overall, subjects reported low stimulation sensation intensities, with only a few instances of very uncomfortable or painful sensations (Figure 8). The topical BL10 anesthetic gel effectively reduced self-reported sensations in the novel tDCS+BL10 condition, as indicated by lower average intensity ratings compared to the tDCS group and even the sham group. However, due to the overall low reported intensities, these differences were only significant in the last session, both compared to the tDCS (p_tDCS – tDCS+BL10_ = 0.0019) and sham group (p_tDCS+BL10 – Sham_ = 0.0008).

**Figure 8.**
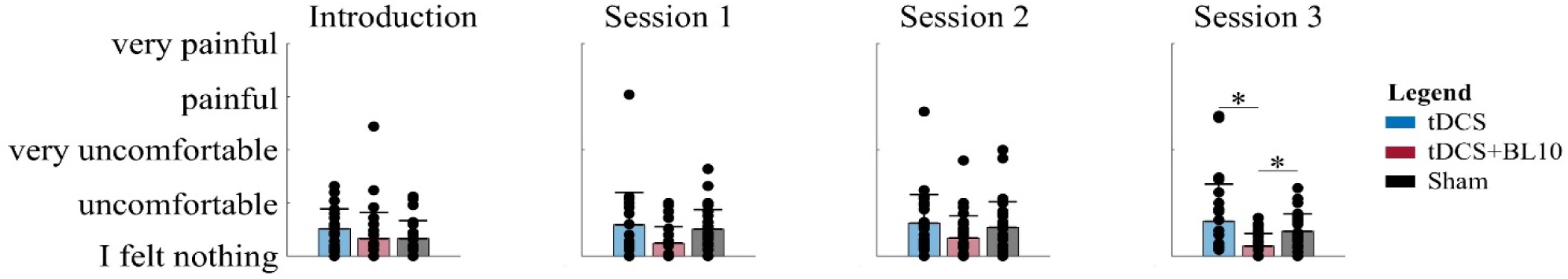
**Stimulation sensations**. Bar plots indicating the mean self-reported stimulation sensations per stimulation condition: tDCS (blue), tDCS+BL10 (red), and sham (black) for the introduction session, session 1, session 2, and session 3, respectively, with error bars indicating standard deviation. On average, the stimulation sensations were lower in the tDCS+BL10 condition, where topical anesthetics under the electrodes reduced scalp sensations, but this difference was only significant (*) in session 3.

### Side effects

Overall, most subjects reported few to no side effects, with the most reported being sensations in the scalp including tingling, itchiness, pinching, warm and slightly burning sensations (Figure 9). Although frequently reported, most subjects rated these side effects as very slightly noticeable, but intensity levels varied significantly with stimulation condition (ANOVA main effect of stimulation condition: p_tingling_ < 0.0001, p_itchiness_ < 0.0001, p_pinching_ < 0.0001, and p_burning_ = 0.0035), expect for warm sensations (ANOVA main effect of stimulation condition: p_warm_ = 0.7514), with the detailed post-hoc comparisons provided in the Supplementary Materials.

**Figure 9.**
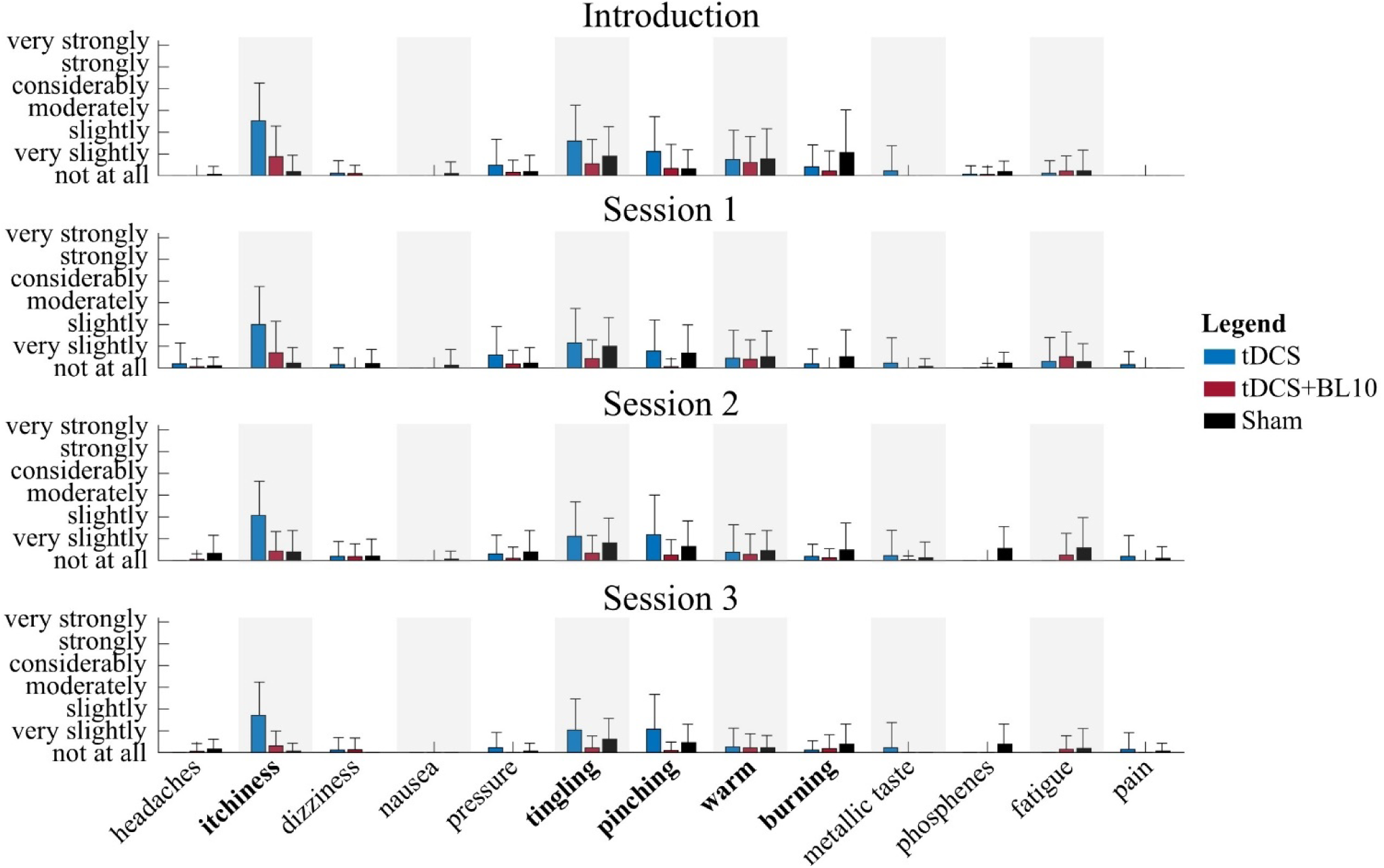
Side effects. Average intensity of side effects during tDCS (blue), tDCS+BL10 (red), and sham (black) stimulation across sessions. Tingling, itchiness, pinching, warm, and burning sensations (bolt) were most reported, whereas other side effects such as headaches, dizziness, nausea, pressure on the scalp, metallic taste, phosphenes, fatigue, and pain were reported much less frequently.

In addition to the significant differences in intensity levels, notable differences were also observed in the occurrence of side effects. Most subjects in the sham condition reported experiencing side effects only once at the onset of stimulation (Table S3), whereas subjects in the tDCS condition reported experiencing side effects at multiple occasions or continuously throughout stimulation (Table S1). Subjects in the tDCS+BL10 condition generally reported minimal side effects (Figure S2, Table S3).

## Discussion

In this study we aimed to replicate the effect of 1mA anodal tDCS over M1 on motor sequence learning in a multiple session serial reaction time task paradigm to determine whether tDCS’s neuromodulatory effects are driven by the resulting electric field in the cortex, as originally hypothesized (Nitsche and Paulus 2000), or by an increase in cortical excitability resulting from stimulation of peripheral nerves in the scalp mechanism (van Boekholdt et al. 2021). Despite some earlier concerns regarding replicability (Horvath et al. 2016; Keitel et al. 2018) and uncertainty about the exact timing of tDCS effects on motor sequence learning, whether they occur during practice and concurrent stimulation (Nitsche et al. 2003; Stagg et al. 2011), immediately afterwards (Dumel et al. 2016), after 24h consolidation (Kang and Paik 2011), or after multiple sessions (Buch et al. 2017; Reis et al. 2009), we hypothesized that a significant effect would be evident after three sessions (Hashemirad et al. 2016).

Contrary to our expectations, we found no evidence that 1 mA anodal tDCS improves motor sequence learning, not during practice and concurrent stimulation, nor immediately following the first practice session, after three sessions, or after consolidation. Hence, we were unable to investigate our main hypothesis regarding the underlying neurophysiological mechanisms and determine the relative contributions of the transcranial and the transcutaneous tDCS mechanisms in mediating behavioral effects. Although the tDCS literature generally proposed motor learning as among the most reliable and consistent behavioral outcomes (Buch et al. 2017) when we designed the experiment, and despite earlier studies showing significant effects of tDCS on motor sequence learning (Dumel et al. 2016; Kang and Paik 2011; Liebrand et al. 2020; Nitsche et al. 2003; Shilo and Lavidor 2019; Stagg et al. 2011), several other groups have failed to replicate these findings (Horvath et al. 2016; Hsu et al. 2025; Keitel et al. 2018; Shilo and Lavidor 2019).

For our study, we applied stimulation parameters previously shown to facilitate motor sequence learning (Buch et al. 2017; Hashemirad et al. 2016; Reis et al. 2009). However, some might argue that the applied stimulation amplitude of 1 mA was insufficient to elicit significant effects, as another study was able to demonstrate online effects of tDCS on motor learning using higher stimulation amplitudes (Hsu et al. 2023). Yet, many other studies highlighted that increasing the stimulation amplitude does not necessarily increase tDCS’s efficacy (Khalil, Karim, and Godde 2023), and sometimes even reversed observed effects (Agboada et al. 2019). Another important aspect to point out in this discussion is the motor sequence learning paradigm in our study. Our results indicated that all stimulation groups exhibited sustained improvements in motor sequence learning, both within each practice session, as well as across the sessions. This means that subjects had not yet reached their maximal performance after two practice sessions yet continued to improve until the end of the experiment. This demonstrates the sensitivity of our motor sequence learning paradigm, as it provided the potential for tDCS to further enhance learning in each session. Nevertheless, no significant differences were observed between the stimulation conditions.

A final aspect of the experimental setup in our study that is important to highlight in this discussion is the use of our newly developed tDCS cup electrodes (Kerstens et al. 2025) rather than conventional rubber patch electrodes. Given their properties, accurately positioning rubber patch electrodes at the desired location and maintaining that position throughout stimulation can be challenging. We therefore developed new tDCS electrodes designed to enhance precision, reproducibility, and reliability of tDCS in humans by addressing commonly reported limitations of conventional electrodes. An important design consideration was to improve conductivity at the electrode-scalp interface, which can be highly variable with conventional electrodes due to factors such as gel leakage, unintended electrical bridging, or electrode displacement during stimulation (Kerstens et al. 2025). However, by achieving a lower impedance at the electrode-scalp interface, with a resistivity of approximately ρ = 0.3 Ωm (Kerstens et al. 2022) using our newly developed tDCS cup electrodes, the voltage required to sustain a constant direct current is reduced. This may have had a potentially greater impact on the observed tDCS outcomes than originally anticipated. It is plausible that the effects of tDCS depend more on the resulting electric potential than on the applied current, which in turn is directly correlated to the impedance at the electrode–scalp interface. Surprisingly, this alternative hypothesis has been overlooked by the tDCS field, even though intuitively the externally applied potential is much more likely to have a proportional impact on the resulting electric field than the applied current. It might even explain why some studies have found that higher stimulation amplitudes did not necessarily induce more profound effects (Khalil et al. 2023). If observed tDCS effects are primarily driven by the resulting voltage rather than the applied current, using our newly developed cup electrodes instead of conventional patch electrodes may have inadvertently obscured a potential effect. This may explain why our study failed to replicate earlier findings, whereas other studies using a similar approach with conventional tDCS electrodes were able to demonstrate significant effects on motor sequence learning (Greeley et al. 2020; Hsu et al. 2023).

These conflicting findings further underscore the challenges of reproducibility and reliability in the tDCS field (Héroux et al. 2017; Minarik et al. 2016). Importantly, this issue extends not only to behavioral outcomes but also to the replication of the most fundamental physiological effects. In the seminal study from Nitsche and Paulus in 2000 (Nitsche and Paulus 2000), anodal tDCS over the motor cortex elicited a prolonged increase in cortical excitability as measured by TMS motor evoked potentials. Interestingly, however, recent double-blind placebo-controlled studies failed to replicate these results (Hsu et al. 2025; Jonker et al. 2021; Lescrauwaet et al. 2024), showing no effect of tDCS on cortical excitability in a much larger sample size. To determine whether the absence of a significant effect could be explained by factors predicting individual responsiveness to tDCS, they further investigated whether individual differences in two previously identified variables could classify subjects as tDCS-responders or non-responders. Previous research has proposed that the efficiency of early indirect wave recruitment in the motor cortex, measured by the latency difference between anterior-posterior and later-medial TMS pulses (Hamada et al. 2013), may predict responsiveness to tDCS (Davidson, Bolic, and Tremblay 2016; Wiethoff, Hamada, and Rothwell 2014). Other studies have identified that carriers of brain-derived neurotrophic factor (BDNF) Val66Met polymorphism (rs6265) show a decrease in the amount of BDNF that is released from activated cells (Shah-Basak et al. 2020) and are therefore more susceptible to enhanced cortical excitability in response to anodal tDCS compared to non-carriers (Antal et al. 2010; Puri et al. 2015). Nevertheless, after performing an exploratory analysis to investigate the predictive power of these two factors in predicting individual responsiveness to tDCS, Jonkers et al. found no evidence for the existence of tDCS responders and non-responders (Jonker et al. 2021) and highlighted that previous studies did not test for subpopulations but identified rather arbitrary thresholds to classify subjects as tDCS-responders or non-responders (Chew, Ho, and Loo 2015; López-Alonso et al. 2015; Tremblay et al. 2016).

Although we aimed to investigate the neurophysiological mechanisms underlying the effect of tDCS (van Boekholdt et al. 2021), we were unable to test our hypothesis because we could not replicate the effect of tDCS on motor sequence learning in the first place. While some of the variability observed in behavioral tDCS studies may be attributed to differences in study paradigms, stimulation parameters, or application methods, it is particularly concerning that multiple recent studies have failed to replicate (Hsu et al. 2025; Jonker et al. 2021; Lescrauwaet et al. 2024) the seminal findings of Nitsche and Paulus (Nitsche and Paulus 2000). Considering the growing number of non-replication studies, we believe it is important to reinitiate the discussion on the reliability of tDCS as a neuromodulation technique (Héroux et al. 2017; Minarik et al. 2016). Instead of exploring more clinical applications, we recommend focusing first on fundamental research aiming at systematically investigating the neurophysiological mechanisms of tDCS in animal studies using our free-moving rat model (van Boekholdt et al. 2023). After identifying if tDCS effects in animals are primarily caused by the transcranial or transcutaneous mechanism, our novel tDCS control condition (Kerstens et al. 2022) that blocks the transcutaneous tDCS mechanism during stimulation could be used to investigate if these fundamental findings also translate to humans.

## Conclusion

In the present study, we aimed to investigate the neurophysiological mechanisms underlying observed behavioral effects of tDCS. Our results confirmed the sensitivity of our motor sequence learning paradigm and confirmed it provided the potential for tDCS to further enhance learning each session. Nevertheless, we found no significant effect of tDCS and were therefore unable to test our main hypothesis regarding the underlying neurophysiological mechanisms. These non-replication of one of the presumably most reliable tDCS effects in a much larger sample size than the original studies, among other similar studies reporting non-replications of previous findings, should reinitiate the discussion regarding the efficacy of tDCS as a neuromodulation technique. Future work should focus on fundamental research to investigate if tDCS effects are driven by the resulting electric field in the cortex or if it elicits its neuromodulatory effects by stimulation of peripheral nerves in the scalp, as a deeper understanding of the mechanisms underlying tDCS effects is essential for optimizing the technique and establishing it as a reliable tool for clinical neuromodulation applications in the future.

## Data Availability

All data produced in the present study are available upon reasonable request to the authors.

## Acknowledgments

We would like to thank all our subjects for participating in our study.

## Author Contributions

The authors’ contributions were as follows. Silke Kerstens: hypothesis, methodology, experimental design, paradigm optimization, design and programming of scripts for data acquisition, data acquisition, study administration, data analysis and manuscript writing. Luuk van Boekholdt: methodology, experimental design, data acquisition. Hans Vanderheyden and Louise De Smedt: data acquisition. Nina Seminck and Tine Van Bogaert: support in pilot data collection. Genevieve Albouy and Bradley Ross King: methodology, experimental design, scripts for data acquisition, manuscript review. Jean-Jacques Orban de Xivry: methodology and experimental design. Myles Mc Laughlin: hypothesis, methodology and experimental design.

## References

Agboada, Desmond, Mohsen Mosayebi Samani, Asif Jamil, Min Fang Kuo, and Michael A. Nitsche. 2019. “Expanding the Parameter Space of Anodal Transcranial Direct Current Stimulation of the Primary Motor Cortex.” Scientific Reports 9(1). doi:10.1038/s41598-019-54621-0.

Antal, Andrea, Leila Chaieb, Vera Moliadze, Katia Monte-Silva, Csaba Poreisz, Nivethida Thirugnanasambandam, Michael A. Nitsche, Moneef Shoukier, Harald Ludwig, and Walter Paulus. 2010. “Brain-Derived Neurotrophic Factor (BDNF) Gene Polymorphisms Shape Cortical Plasticity in Humans.” Brain Stimulation 3(4):230–37. doi:10.1016/j.brs.2009.12.003.

Asamoah, Boateng, Ahamd Khatoun, and Myles Mc Laughlin. 2019. “TACS Motor System Effects Can Be Caused by Transcutaneous Stimulation of Peripheral Nerves.” Nature Communications 10(1):266. doi:10.1038/s41467-018-08183-w.

Basner, Mathias, and Joshua Rubinstein. 2011. “Fitness for Duty: A 3-Minute Version of the Psychomotor Vigilance Test Predicts Fatigue-Related Declines in Luggage-Screening Performance.” Journal of Occupational and Environmental Medicine 53(10):1146–54. doi:10.1097/JOM.0b013e31822b8356.

van Boekholdt, Luuk, Silke Kerstens, Kaydee Decloedt, and Myles Mc Laughlin. 2023. “A Novel Free-Moving Rat Model of Transcranial Direct Current Stimulation.” Brain Stimulation 16(6):1601–3. doi: 10.1016/j.brs.2023.10.010.

van Boekholdt, Luuk, Silke Kerstens, Ahmad Khatoun, Boateng Asamoah, and Myles Mc Laughlin. 2021. “TDCS Peripheral Nerve Stimulation: A Neglected Mode of Action?” Molecular Psychiatry 26(2):456–61. doi:10.1038/s41380-020-00962-6.

Brunoni, Andre Russowsky, Joao Amadera, Bruna Berbel, Magdalena Sarah Volz, Brenno Gomes Rizzerio, and Felipe Fregni. 2011. “A Systematic Review on Reporting and Assessment of Adverse Effects Associated with Transcranial Direct Current Stimulation.” International Journal of Neuropsychopharmacology 14(8):1133–45. doi: 10.1017/S1461145710001690.

Buch, Ethan R., Emiliano Santarnecchi, Andrea Antal, Jan Born, Pablo A. Celnik, Joseph Classen, Christian Gerloff, Mark Hallett, Friedhelm C. Hummel, Michael A. Nitsche, Alvaro Pascual-Leone, Walter J. Paulus, Janine Reis, Edwin M. Robertson, John C. Rothwell, Marco Sandrini, Heidi M. Schambra, Eric M. Wassermann, Ulf Ziemann, and Leonardo G. Cohen. 2017. “Effects of TDCS on Motor Learning and Memory Formation: A Consensus and Critical Position Paper.” Clinical Neurophysiology 128(4):589–603. doi:10.1016/j.clinph.2017.01.004.

Chew, Taariq, Kerrie-Anne Ho, and Colleen K. Loo. 2015. “Inter-and Intra-Individual Variability in Response to Transcranial Direct Current Stimulation (TDCS) at Varying Current Intensities.” *Brain Stimulation: Basic*, Translational, and Clinical Research in Neuromodulation 8(6):1130–37. doi:10.1016/j.brs.2015.07.031.

Davidson, Travis W., Miodrag Bolic, and François Tremblay. 2016. “Predicting Modulation in Corticomotor Excitability and in Transcallosal Inhibition in Response to Anodal Transcranial Direct Current Stimulation.” Frontiers in Human Neuroscience 10(FEB2016). doi:10.3389/fnhum.2016.00049.

Dinges, David F., and John W. Powell. 1985. “Microcomputer Analyses of Performance on a Portable, Simple Visual RT Task during Sustained Operations.” Behavior Research Methods, Instruments, & Computers 17(6):652–55. doi:10.3758/BF03200977.

Dumel, G., M. E. Bourassa, M. Desjardins, N. Voarino, C. Charlebois-Plante, J. Doyon, and Louis De Beaumont. 2016. “Multisession Anodal TDCS Protocol Improves Motor System Function in an Aging Population.” Neural Plasticity 2016:15–18. doi:10.1155/2016/5961362.

Fanselow, Erika E. 2012. “Central Mechanisms of Cranial Nerve Stimulation for Epilepsy.” Surgical Neurology International 3(Suppl 4):S247–54. doi:10.4103/2152-7806.103014.

Greeley, Brian, Jonathan S. Barnhoorn, Willem B. Verwey, and Rachael D. Seidler. 2020. “Multi-Session Transcranial Direct Current Stimulation Over Primary Motor Cortex Facilitates Sequence Learning, Chunking, and One Year Retention.” Frontiers in Human Neuroscience 14(March):1–18. doi:10.3389/fnhum.2020.00075.

Hamada, Masashi, Nagako Murase, Alkomiet Hasan, Michelle Balaratnam, and John C. Rothwell. 2013. “The Role of Interneuron Networks in Driving Human Motor Cortical Plasticity.” Cerebral Cortex 23(7):1593–1605. doi:10.1093/cercor/bhs147.

Hashemirad, Fahimeh, Maryam Zoghi, Paul B. Fitzgerald, and Shapour Jaberzadeh. 2016. “The Effect of Anodal Transcranial Direct Current Stimulation on Motor Sequence Learning in Healthy Individuals: A Systematic Review and Meta-Analysis.” Brain and Cognition 102:1–12. doi:10.1016/j.bandc.2015.11.005.

Héroux, Martin E., Colleen K. Loo, Janet L. Taylor, and Simon C. Gandevia. 2017. “Questionable Science and Reproducibility in Electrical Brain Stimulation Research” edited by J. M. Wicherts. PLOS ONE 12(4):e0175635. doi:10.1371/journal.pone.0175635.

Horvath, Jared Cooney, Olivia Carter, and Jason D. Forte. 2016. “No Significant Effect of Transcranial Direct Current Stimulation (TDCS) Found on Simple Motor Reaction Time Comparing 15 Different Simulation Protocols.” Neuropsychologia 91:544–52. doi:10.1016/j.neuropsychologia.2016.09.017.

Hsu, Gavin, Zhenous Hadi Jafari, Abdelrahman Ahmed, Dylan J. Edwards, Leonardo G. Cohen, and Lucas C. Parra. 2025. “Dose–Response of TDCS Effects on Motor Learning and Cortical Excitability: A Preregistered Study.” Imaging Neuroscience 3. doi:10.1162/imag_a_00431.

Hsu, Gavin, A. Duke Shereen, Leonardo G. Cohen, and Lucas C. Parra. 2023. “Robust Enhancement of Motor Sequence Learning with 4 MA Transcranial Electric Stimulation.” Brain Stimulation 16(1):56–67. doi:10.1016/j.brs.2022.12.011.

Jonker, Zeb D., Carolin Gaiser, Joke H. M. Tulen, Gerard M. Ribbers, Maarten A. Frens, and Ruud W. Selles. 2021. “No Effect of Anodal TDCS on Motor Cortical Excitability and No Evidence for Responders in a Large Double-Blind Placebo-Controlled Trial.” Brain Stimulation 14(1):100–109. doi:10.1016/j.brs.2020.11.005.

Kang, Eun K., and Nam Jong Paik. 2011. “Effect of a TDCS Electrode Montage on Implicit Motor Sequence Learning in Healthy Subjects.” Experimental and Translational Stroke Medicine 3(1):4. doi:10.1186/2040-7378-3-4.

Keitel, Ariane, Henning Øfsteng, Vanessa Krause, and Bettina Pollok. 2018. “Anodal Transcranial Direct Current Stimulation (TDCS) over the Right Primary Motor Cortex (M1) Impairs Implicit Motor Sequence Learning of the Ipsilateral Hand.” Frontiers in Human Neuroscience 12(July):1–7. doi:10.3389/fnhum.2018.00289.

Kerstens, Silke, Luuk van Boekholdt, Jean Jacques Orban de Xivry, and Myles Mc Laughlin. 2025. “3D-Printable Stimulation Electrodes to Improve Precision, Reproducibility, and Reliability of Transcranial Electric Current Stimulation.” Clinical Neurophysiology 179. doi:10.1016/j.clinph.2025.2110981.

Kerstens, Silke, Jean-Jacques Orban de Xivry, and Myles Mc Laughlin. 2022. “A Novel TDCS Control Condition Using Optimized Anesthetic Gel to Block Peripheral Nerve Input.” Frontiers in Neurology 13. doi:10.3389/fneur.2022.1049409.

Kessler SK, Turkeltaub PE, Benson JG, Hamilton RH. 2012. “Differences in the Experience of Active and Sham Transcranial Direct Current Stimulation.” Brain Stimulation. doi:10.1016/j.brs.2011.02.007.

Khalil, Radwa, Ahmed A. Karim, and Ben Godde. 2023. “Less Might Be More: 1 MA but Not 1.5 MA of TDCS Improves Tactile Orientation Discrimination.” IBRO Neuroscience Reports 15:186–92. doi:10.1016/j.ibneur.2023.08.003.

Lescrauwaet, Emma, Mathieu Sprengers, Evelien Carrette, Chloé Algoet, Ann Mertens, Debby Klooster, Steven Beumer, Rob Mestrom, Robrecht Raedt, Paul Boon, and Kristl Vonck. 2024. “Investigating the Working Mechanism of Transcranial Direct Current Stimulation.” Neuromodulation. doi:10.1016/j.neurom.2024.05.002.

Liebrand, Matthias, Anke Karabanov, Daria Antonenko, Agnes Flöel, Hartwig R. Siebner, Joseph Classen, Ulrike M. Krämer, and Elinor Tzvi. 2020. “Beneficial Effects of Cerebellar TDCS on Motor Learning Are Associated with Altered Putamen-Cerebellar Connectivity: A Simultaneous TDCS-FMRI Study.” NeuroImage 223(April):117363. doi:10.1016/j.neuroimage.2020.117363.

Loh, Sylvia, Nicole Lamond, Jill Dorrian, Gregory Roach, and Drew Dawson. 2004. “The Validity of Psychomotor Vigilance Tasks of Less than 10-Minute Duration.” *Behavior Research Methods*, Instruments, and Computers 36(2):339–46. doi:10.3758/BF03195580.

López-Alonso, Virginia, Miguel Fernández-del-Olmo, Alessia Costantini, Juan Jose Gonzalez-Henriquez, and Binith Cheeran. 2015. “Intra-Individual Variability in the Response to Anodal Transcranial Direct Current Stimulation.” Clinical Neurophysiology 126(12):2342–47. doi:10.1016/j.clinph.2015.03.022.

Matsangas, Panagiotis, and Nita Lewis Shattuck. 2018. “Agreement between the 3-Minute Psychomotor Vigilance Task (PVT) Embedded in a Wrist-Worn Device and the Laptop-Based PVT.” Proceedings of the Human Factors and Ergonomics Society 2:666–70. doi:10.1177/1541931218621151.

Mercante, Beniamina, Francesca Ginatempo, Andrea Manca, Francesco Melis, Paolo Enrico, and Franca Deriu. 2018. “Anatomo-Physiologic Basis for Auricular Stimulation.” Medical Acupuncture 30(3):141–50. doi:10.1089/acu.2017.1254.

Minarik, Tamas, Barbara Berger, Laura Althaus, Veronika Bader, Bianca Biebl, Franziska Brotzeller, Theodor Fusban, Jessica Hegemann, Lea Jesteadt, Lukas Kalweit, Miriam Leitner, Francesca Linke, Natalia Nabielska, Thomas Reiter, Daniela Schmitt, Alexander Spraetz, and Paul Sauseng. 2016. “The Importance of Sample Size for Reproducibility of TDCS Effects.” Frontiers in Human Neuroscience 10:453. doi:10.3389/fnhum.2016.00453.

Nitsche, M. A., and W. Paulus. 2000. “Excitability Changes Induced in the Human Motor Cortex by Weak Transcranial Direct Current Stimulation.” The Journal of Physiology 527(3):633–39. doi: 10.1111/j.1469-7793.2000.t01-1-00633.x

Nitsche, Michael A., Astrid Schauenburg, Nicolas Lang, David Liebetanz, Cornelia Exner, Walter Paulus, and Frithjof Tergau. 2003. “Facilitation of Implicit Motor Learning by Weak Transcranial Direct Current Stimulation of the Primary Motor Cortex in the Human.” Journal of Cognitive Neuroscience 15(4):619–26. doi:10.1162/089892903321662994.

Poreisz, Csaba, Klára Boros, Andrea Antal, and Walter Paulus. 2007. “Safety Aspects of Transcranial Direct Current Stimulation Concerning Healthy Subjects and Patients.” Brain Research Bulletin 72(4–6):208–14. doi:10.1016/j.brainresbull.2007.01.004.

Puri, Rohan, Mark R. Hinder, Hakuei Fujiyama, Rapson Gomez, Richard G. Carson, and Jeffery J. Summers. 2015. “Duration-Dependent Effects of the BDNF Val66Met Polymorphism on Anodal TDCS Induced Motor Cortex Plasticity in Older Adults: A Group and Individual Perspective.” Frontiers in Aging Neuroscience 7(JUN). doi:10.3389/fnagi.2015.00107.

Radman, Thomas, Raddy L. Ramos, Joshua C. Brumberg, and Marom Bikson. 2009. “Role of Cortical Cell Type and Morphology in Subthreshold and Suprathreshold Uniform Electric Field Stimulation.” Brain Stimulation 2(4):215–28. doi:10.1016/j.brs.2009.03.007.Role.

Rampersad, Sumientra M., Arno M. Janssen, Felix Lucka, Umit Aydin, Benjamin Lanfer, Seok Lew, Carsten H. Wolters, Dick F. Stegeman, and Thom F. Oostendorp. 2014. “Simulating Transcranial Direct Current Stimulation With a Detailed Anisotropic Human Head Model.” IEEE Transactions on Neural Systems and Rehabilitation Engineering 22(3):441–52. doi:10.1109/TNSRE.2014.2308997.

Reis, Janine, Heidi M. Schambra, Leonardo G. Cohen, Ethan R. Buch, Brita Fritsch, Eric Zarahn, Pablo A. Celnik, and John W. Krakauer. 2009. “Noninvasive Cortical Stimulation Enhances Motor Skill Acquisition over Multiple Days through an Effect on Consolidation.” Proceedings of the National Academy of Sciences of the United States of America 106(5):1590–95. doi:10.1073/pnas.0805413106.

Van Roy, Anke, Geneviève Albouy, Ryan D. Burns, and Bradley R. King. 2024. “Children Exhibit a Developmental Advantage in the Offline Processing of a Learned Motor Sequence.” Communications Psychology 2(1). doi:10.1038/s44271-024-00082-9.

Shah-Basak P, Harvey DY, Parchure S, Faseyitan O, Sacchetti D, Ahmed A, Thiam A, Lohoff FW, Hamilton RH. Brain-Derived Neurotrophic Factor Polymorphism Influences Response to Single-Pulse Transcranial Magnetic Stimulation at Rest. Neuromodulation. 2020 Oct 9:10.1111/ner.13287. doi: 10.1111/ner.13287. Epub ahead of print. PMID: 33090650; PMCID: PMC8032803.

Shilo, Gali, and Michal Lavidor. 2019. “Non-Linear Effects of Cathodal Transcranial Direct Current Stimulation (TDCS) of the Primary Motor Cortex on Implicit Motor Learning.” Experimental Brain Research 237(4):919–25. doi:10.1007/s00221-019-05477-3.

So, P. P. M., M. A. Stuchly, and J. A. Nyenhuis. 2004. “Peripheral Nerve Stimulation by Gradient Switching Fields in Magnetic Resonance Imaging.” IEEE Transactions on Biomedical Engineering 51(11):1907–14. doi:10.1109/TBME.2004.834251.

Stagg, C. J., G. Jayaram, D. Pastor, Z. T. Kincses, P. M. Matthews, and H. Johansen-Berg. 2011. “Polarity and Timing-Dependent Effects of Transcranial Direct Current Stimulation in Explicit Motor Learning.” Neuropsychologia 49(5):800–804. doi:10.1016/j.neuropsychologia.2011.02.009.

Tremblay, Sara, Félix Larochelle-Brunet, Louis Philippe Lafleur, Sofia El Mouderrib, Jean François Lepage, and Hugo Théoret. 2016. “Systematic Assessment of Duration and Intensity of Anodal Transcranial Direct Current Stimulation on Primary Motor Cortex Excitability.” European Journal of Neuroscience 44(5):2184–90. doi:10.1111/ejn.13321.

Wiethoff, Sarah, Masashi Hamada, and John C. Rothwell. 2014. “Variability in Response to Transcranial Direct Current Stimulation of the Motor Cortex.” *Brain Stimulation: Basic*, Translational, and Clinical Research in Neuromodulation 7(3):468–75. doi:10.1016/j.brs.2014.02.003.

