## Supplemental Materials for "Does anodal tDCS over M1 really enhance motor sequence learning? A non-replication of earlier findings in a double-blind, pre-registered large-sample study in humans"

### Supplementary Materials

#### Side effects

*Post-hoc analyses*

Post-hoc analyses revealed that subjects in the tDCS condition reported significantly higher intensities for itchiness compared to sham in all sessions (introduction session: $p_{itchiness} < 0.0001$, session 1: $p_{itchiness} = 0.0031$, session 2: $p_{itchiness} = 0.0001$, session 3: $p_{itchiness} = 0.0096$), but not for tingling, pinching and burning sensations in the scalp (introduction session: $p_{tingling} = 0.0854$, $p_{burning} = 0.1231$, session 1: $p_{tingling} = 0.6990$, $p_{pinching} = 0.7801$, $p_{burning} = 0.2211$, session 2: $p_{tingling} = 0.3982$, $p_{pinching} = 0.1789$, $p_{burning} = 0.2520$, session 3: $p_{tingling} = 0.1839$, $p_{pinching} = 0.0638$, $p_{burning} = 0.1577$), with the exception of pinching sensation in the scalp in the introduction session ($p_{pinching} = 0.0212$).

Similarly, subjects in the tDCS condition reported significantly higher intensities for itchiness, tingling and pinching sensations in the scalp compared to the tDCS+BL10 condition in all sessions (introduction session: $p_{tingling} = 0.0050$, $p_{itchiness} = 0.0002$ , $p_{pinching} = 0.0306$, session 1: $p_{tingling} = 0.0285$, $p_{itchiness} = 0.0025$, $p_{pinching} = 0.0069$, session 2: $p_{tingling} = 0.0168$, $p_{itchiness} < 0.0001$, $p_{pinching} = 0.0082$, session 3: $p_{tingling} = 0.0033$, $p_{itchiness} < 0.0001$, $p_{pinching} = 0.0011$) but not for burning sensation in the scalp (introduction session: $p_{burning} = 0.4387$, session 1: $p_{burning} = 0.1232$, session 2: $p_{burning} = 0.6125$, session 3: $p_{burning} = 0.6225$). Whereas the differences between the reported intensities for the side effects between the tDCS+BL10 and sham condition were more variable. Some were significant (introduction session: $p_{burning} = 0.0286$, session 1: $p_{tingling} = 0.0418$, $p_{pinching} = 0.0109$, $p_{burning} = 0.0193$, session 3: $p_{tingling} = 0.0419$, $p_{itchiness} = 0.0408$, $p_{pinching} = 0.0308$,), while others were not (introduction session: $p_{tingling} =0.2520$, $p_{itchiness} = 0.1831$, $p_{pinching} = 0.9659$, session 1: $p_{itchiness} = 0.7411$, session 2: $p_{tingling} = 0.0595$, $p_{itchiness} = 0.4280$, $p_{pinching} = 0.0986$, , $p_{burning} = 0.1159$, session 3: $p_{burning} = 0.3002$).

*Side effect occurrence*

**Table S1** Occurrence of reported side effects during stimulation for tDCS

| | Side effect | Once | | | | At beginning | | | | At the end | | | | Multiple times | | | | Consistently | | | |
|---|---|---|---|---|---|---|---|---|---|---|---|---|---|---|---|---|---|---|---|---|---|
| | | S0 | S1 | S2 | S3 | S0 | S1 | S2 | S3 | S0 | S1 | S2 | S3 | S0 | S1 | S2 | S3 | S0 | S1 | S2 | S3 |
| Skin sens. | Itchiness | 1 | 1 | 1 | 1 | 3 | 1 | 3 | 1 | | 1 | 1 | | 7 | 9 | 9 | 8 | 10 | 7 | 7 | 8 |
| | Tingling | 2 | 1 | 1 | 2 | 5 | 4 | 4 | 2 | | | | | 7 | 2 | 3 | 3 | 2 | 4 | 3 | 4 |
| | Pinching | 1 | 1 | 1 | 1 | 3 | 2 | 4 | 3 | | | | | 2 | 1 | 2 | 3 | 4 | 4 | 4 | 4 |
| | Warm | 2 | 1 | 1 | | | 2 | | 1 | 1 | | | | 1 | | | 1 | 3 | 1 | 2 | 1 |
| | Burning | | | | | 3 | 1 | 2 | 2 | 1 | | | | 1 | 1 | 1 | | | | | |
| Other side effects | Headaches | | | | | | | | | | | | | | | | 1 | | 2 | | |
| | Pressure | | | | | | | | | | | | | 1 | 2 | 1 | | 3 | 3 | 2 | 3 |
| | Dizziness | | | 1 | | | | | | | | | | | | | 1 | 1 | 1 | 1 | |
| | Nausea | | | | | | | | | | | | | | | | | | | | |
| | Metallic | | | | | | | | | | | | | | | | | 1 | 1 | 1 | 1 |
| | Phosphene | | | | | 1 | | | | | | | | | | | | | | | |
| | Fatigue | | | | | | | | | | | | | | 2 | | | 1 | 1 | | |
| | Pain | | | | | | 2 | 1 | 1 | | | | | | | | | | | | |

Note: The numbers in the columns indicate the number of subjects that indicated they experienced a particular side effect either only once at the onset of stimulation (at beginning), or once at the end of stimulation (at the end), or once at any other time during stimulation (once), at multiple occasions during stimulation (multiple times) or consistently throughout stimulation for the introduction session (S0), session 1 (S1), session 2 (S2) and session 3 (S3) respectively per side effect.

**Table S2** Occurrence of reported side effects during stimulation for tDCS+BL10

| | Side effect | Once | | | | At beginning | | | | At the end | | | | Multiple times | | | | Consistently | | | |
|---|---|---|---|---|---|---|---|---|---|---|---|---|---|---|---|---|---|---|---|---|---|
| | | S0 | S1 | S2 | S3 | S0 | S1 | S2 | S3 | S0 | S1 | S2 | S3 | S0 | S1 | S2 | S3 | S0 | S1 | S2 | S3 |
| Skin sens. | Itchiness | 4 | 1 | 1 | 1 | 2 | 1 | | 2 | 1 | | | | 3 | 4 | 4 | 2 | 2 | 2 | 2 | 1 |
| | Tingling | 1 | 1 | 2 | | 5 | 3 | 2 | 5 | | | 1 | | | 3 | 1 | | 2 | | | |
| | Pinching | 1 | 1 | | | 2 | | 4 | 2 | | | | | 1 | | | | | | | |
| | Warm | 2 | 1 | 1 | 1 | | 1 | | 1 | | | | | 1 | 2 | 1 | 2 | 6 | 3 | 2 | |
| | Burning | | | | | | | 2 | 2 | | | | | 1 | | | 1 | 1 | | 1 | |
| Other side effects | Headaches | | | 1 | | | | | | | | | | | 1 | 1 | 1 | | | | |
| | Pressure | | | | | 2 | 1 | | | | | | | | | | | 1 | 2 | 1 | |
| | Dizziness | | | 1 | | 1 | | | | | | 1 | | 1 | | 1 | 2 | | | | |
| | Nausea | | | | | | | | | | | | | | | | | | | | |
| | Metallic | | | 1 | | | | | | | | | | | | | | | | | |
| | Phosphene | 1 | | | | | 1 | | | | | | | | | | | | | | |
| | Fatigue | 1 | 2 | 1 | | | | | 1 | 2 | 2 | 1 | | | 2 | | 1 | | | | |
| | Pain | | | | | | | | | | | | | | | | | | | | |

Note: The numbers in the columns indicate the number of subjects that indicated they experienced a particular side effect either only once at the onset of stimulation (at beginning), or once at the end of stimulation (at the end), or once at any other time during stimulation (once), at multiple occasions during stimulation (multiple times) or consistently throughout stimulation for the introduction session (S0), session 1 (S1), session 2 (S2) and session 3 (S3) respectively per side effect.

**Table S3** Occurrence of reported side effects during stimulation for sham

| | Side effect | Once | | | | At beginning | | | | At the end | | | | Multiple times | | | | Consistently | | | |
|---|---|---|---|---|---|---|---|---|---|---|---|---|---|---|---|---|---|---|---|---|---|
| | | S0 | S1 | S2 | S3 | S0 | S1 | S2 | S3 | S0 | S1 | S2 | S3 | S0 | S1 | S2 | S3 | S0 | S1 | S2 | S3 |
| Skin sens. | Itchiness | 1 | | | 3 | 6 | 7 | 8 | 8 | 1 | | | | 1 | 3 | 1 | 1 | | 2 | 1 | 1 |
| | Tingling | 1 | | | | 5 | 12 | 10 | 11 | | | | | 3 | 2 | 1 | | 4 | | 1 | |
| | Pinching | | | | | 4 | 7 | 7 | 7 | | | | | | 1 | | 1 | 1 | | 1 | |
| | Warm | | 2 | | | 6 | 5 | 4 | 5 | | | | | 1 | | 2 | | 2 | | 1 | |
| | Burning | | | | | 7 | 5 | 4 | 4 | | | | | | | 1 | 1 | 1 | 1 | | |
| Other side effects | Headaches | | 1 | 3 | 3 | | 1 | 1 | 1 | | | | | | | | | 1 | | 1 | |
| | Pressure | | | | | 1 | 2 | 1 | 1 | | | | | | 1 | 2 | | 1 | | 2 | |
| | Dizziness | | | | | | 2 | 1 | | | | | | | 1 | | | | | 1 | |
| | Nausea | | | | | | | | | 1 | | 1 | | | 1 | | | | | | |
| | Metallic | | | | | | 1 | | | | | | | | | | | | | 1 | |
| | Phosphene | 1 | | | | 4 | 6 | 9 | 7 | | | | | | | | | | | | |
| | Fatigue | | 1 | | | | | | | | 2 | 1 | 1 | 2 | 1 | 1 | 1 | | | 3 | |
| | Pain | | | | | | | | 1 | | | | | | | 1 | | | | | |

Note: The numbers in the columns indicate the number of subjects that indicated they experienced a particular side effect either only once at the onset of stimulation (at beginning), or once at the end of stimulation (at the end), or once at any other time during stimulation (once), at multiple occasions during stimulation (multiple times) or consistently throughout stimulation for the introduction session (S0), session 1 (S1), session 2 (S2) and session 3 (S3) respectively per side effect.
